# Medication Safety Incidents Associated with the Remote Delivery of Primary Care: A Rapid Review

**DOI:** 10.1101/2022.05.19.22275325

**Authors:** Laura L Gleeson, Barbara Clyne, James W. Barlow, Benedict Ryan, Paul Murphy, Emma Wallace, Aoife De Brún, Lisa Mellon, Marcus Hanratty, Mark Ennis, Alice Holton, Muriel Pate, Ciara Kirke, Michelle Flood, Frank Moriarty

## Abstract

**Background:** The COVID-19 pandemic triggered rapid, fundamental changes in how healthcare is delivered in communities, notably increased remote delivery of primary care. While the impact of these changes on medication safety are not yet fully understood, research conducted before the pandemic may provide evidence for possible consequences. This rapid review examines the published literature on medication safety incidents associated with the remote delivery of primary care.

**Objective(s):** To examine the published literature on medication safety incidents associated with the remote delivery of primary care, with a focus on telemedicine and electronic prescribing.

**Methods:** A rapid review was conducted according to the Cochrane Rapid Reviews Methods Group guidance. An electronic search was carried out on Embase and Medline (via PubMed) using key search terms “medication error”, “electronic prescribing”, “telemedicine” and “primary care”. Identified studies were synthesised narratively; reported medication safety incidents were categorised according to the WHO Conceptual Framework for the International Classification for Patient Safety.

**Results:** Fifteen studies were deemed eligible for inclusion in this review. All fifteen studies reported medication incidents associated with electronic prescribing; no studies were identified that reported medication safety incidents associated with telemedicine. The most commonly reported medication safety incidents were ‘wrong label/instruction’ and ‘wrong dose/strength/frequency’. The frequency of medication safety incidents ranged from 0.89 to 81.98 incidents per 100 electronic prescriptions analysed.

**Conclusions:** To our knowledge, this is the first review to examine the literature on medication safety incidents associated with the remote delivery of primary care. Common incident types associated with electronic prescriptions were identified. There was wide variation in reported frequencies of medication safety incidents associated with electronic prescriptions. A gap in the literature was identified regarding medication safety incidents associated with telemedicine. Further research is required to determine the impact of the COVID-19 pandemic on medication safety in primary care.

## Introduction

Worldwide, the declaration of the COVID-19 pandemic by the World Health Organisation (WHO) in March 2020 led to rapid and fundamental changes in the way that healthcare is delivered.(1) Many of these changes occurred in primary care, defined as any health or social care service provided in the community, to prevent the spread of COVID-19 in healthcare facilities such as general practices (GPs) and community pharmacies.(2) Examples include the expansion of medicines home delivery services by community pharmacies, the implementation of electronic prescribing, and the use of technology to deliver care remotely.(3,4) In Ireland, two significant changes were the increased use of technology to conduct both GP and pharmacist consultations (telemedicine) and the introduction of new legislation to allow electronically-transferred prescriptions (e-prescriptions), removing the need for a paper copy.(5,6) By preventing vulnerable patients from congregating in healthcare settings, these changes have clear benefits in minimising infection risk. However, they also reduce opportunities for traditional face-to-face interaction and informal communication between patients and healthcare professionals about safe medicines use, potentially increasing the risk of medication safety incidents.(7,8)

Despite increased levels of awareness and research over the past two decades, medication safety incidents, defined as any preventable event that may cause or lead to inappropriate medication use or patient harm, remain a leading source of preventable harm worldwide.(9–11) Medication safety incidents can occur at any stage during the prescribing, compounding, dispensing, administration, education, monitoring, and use of medicines.(11) The WHO estimates that 1 in every 300 patients dies due to a preventable medical accident, and that up to 4 in 10 patients suffer an adverse event while receiving primary or ambulatory care.(12) Due to variations in definitions and methods used, there is a lack of consensus in the published literature regarding the rate of medication errors occurring in primary care services. Estimates of dispensing errors in community pharmacies range from 0.04% to 24%, while a 2018 systematic review by Assiri *et al*. found that prevalence estimates of prescribing errors, including potentially inappropriate prescribing, ranged from 2% to 94%.(13–15) It is not yet clear whether reduced face-to-face contact between healthcare professionals and patients since the onset of the COVID-19 pandemic has led to an increased risk of medication safety incidents in primary care. Due to the likelihood of the continued remote delivery of primary care beyond the pandemic, it is important to understand and mitigate these risks, if applicable, as soon as possible.(16) Previous research on this topic conducted before the pandemic may provide helpful evidence on the expected consequences of the increased remote delivery of primary care. Therefore, the aim of this rapid review is to examine the pre-pandemic literature to identify the types and causes of medication safety incidents associated with the remote delivery of primary care via electronic prescribing and telemedicine specifically.

## Methods

Due to the time-sensitive nature of this research topic, a rapid review was conducted in accordance with the interim guidance from the Cochrane Rapid Reviews Group.(17) Compared to a traditional systematic review, abbreviated steps in a rapid review include searching fewer databases, double screening 20% of titles and abstracts, single screening of full texts, and synthesising evidence narratively. The study protocol was registered in advance with the International Prospective Register of Systematic Reviews (PROSPERO): **Reg No. CRD42021258580**. Although there is currently no standardised reporting guideline for rapid reviews, the PRISMA statement was used to guide reporting of this study (**Appendix 1**).(18)

### Search Strategy

An electronic search was carried out using Embase and Medline (via PubMed) using the following key search terms: “medication error”, “electronic prescribing”, “telemedicine” and “primary care”. The search strategy was designed with expert input from a librarian (PM) and a systematic review expert (BC). The full search strategy is available in **Appendix 2**. The search strategy was developed between May and June 2021, and the final search was conducted on June 2^nd^ 2021. The search was restricted to English language articles and articles published since January 2000 to June 2nd 2021.

### Inclusion Criteria

Studies that met the following inclusion criteria were included in the review:

- Any peer-reviewed study reporting primary data, including cohort, cross-sectional, case-control, experimental (e.g. randomised controlled trial), and quasi-experimental (e.g. interrupted time series) studies, case series, or other appropriate study design.
- Investigating the types and/or causes of medication safety incidents (medication errors or near misses) associated with remote delivery of primary care electronic prescribing or telemedicine.
- Published in English
- Published since 2000

The definition of medication safety incident used in the review was *‘any preventable event that may cause or lead to inappropriate medication use or patient harm’*.(19) For the purposes of this review, the term ‘medication safety incident’ encompasses both medication errors (i.e. incidents that reach the patient) and near misses (i.e. incidents that are identified before reaching the patient). The term ‘telemedicine’ refers to the use of technology to deliver healthcare remotely, and the term ‘e-prescribing’ refers to the electronic transfer of prescriptions from a physician to a pharmacist without the need for a paper copy.(16,20)

### Exclusion Criteria

The following exclusion criteria were applied:

- Studies not investigating types and/or causes of medication safety incidents (medication errors or near misses) associated with the remote delivery of primary healthcare (with a specific focus on electronic prescribing or telemedicine)
- Studies investigating potentially inappropriate prescribing only
- Studies conducted in secondary or tertiary care
- Studies not available in the English language
- Individual case reports
- Systematic reviews or protocols
- Conference abstracts
- Studies reporting qualitative data only
- Studies not reporting primary data

### Study Selection

Search results were uploaded to the Covidence programme, which was used to improve efficiency of study screening.(21) In accordance with the Cochrane rapid reviews guidance, a pilot exercise was carried out on 30 titles and abstracts to ensure consistency in the application of inclusion and exclusion criteria. Once the inclusion and exclusion criteria were refined, the identified titles and abstracts were screened for eligibility by the screening team (LG, MF, FM, BR, & JB). In accordance with the guidance, 20% of the titles and abstracts were double-screened, while the remaining abstracts were screened by a single reviewer.(17) The full texts were then screened by one reviewer (LG) with consultation with the wider review group when necessary. Excluded studies were checked by a second reviewer (FM) to confirm the reason for exclusion and reasons for exclusion were documented.

### Data Extraction

The following data were extracted from the included studies into a standardised data collection form: study characteristics (e.g. study author, year, design, setting and country, unit of analysis), medication incidents (e.g. types, causes, outcomes and frequencies), how they were defined (e.g. whether errors and near misses were included and differentiated) and measured (e.g. independent review of prescriptions by researchers versus self-report by pharmacists/technicians) and, where applicable, details of a comparison group and relative risk or statistical test to compare risk or rate of incidents. The data extraction form was piloted with members of the screening team (LG, MF, FM), data extraction then carried out using Microsoft Excel. The data was extracted by a single reviewer (LG) and verified by a second reviewer (FM).

### Outcomes

The main outcome of interest in this review was the types of medication safety incidents (including medication errors and near misses) associated with the remote delivery of primary care. Secondary outcomes of interest were the frequency, causes and outcomes of medication safety incidents associated with the remote delivery of primary healthcare.

### Quality Assessment

Quality assessments were carried out for all included studies using the Critical Skills Appraisal Checklist (CASP Checklist) for cohort studies. This was pre-specified as the tool which would likely accommodate most eligible studies. Irrelevant cohort items were marked as non-applicable for cross-sectional studies. Assessments were carried out by one reviewer (LG), and verified by a second reviewer (FM).

### Data Synthesis

The incident types reported in each study were categorised according to the WHO Conceptual Framework for the International Classification for Patient Safety (ICPS).(22) Incident types that did not fit the framework, e.g. incomplete prescriptions or missing essential information, were recorded as ‘Other’. Events that did not fall under the definition of medication safety incidents used in this review, e.g. supply problems, information written in the wrong field, or missing signatures, were recorded as Quality Related Events (QREs). According to the Cochrane guidance, a narrative synthesis without meta-analysis was carried out.(17) Where possible, and where it was not already reported, an incident rate was calculated for each study using the following formula: # incidents reported/ # prescriptions analysed * 100.

## Results

In total, 3512 records were identified, of which 38 duplicates were removed. Of the 3474 remaining titles, 3411 were excluded during title and abstract screening. Therefore, 63 full texts were assessed for eligibility, with 15 deemed eligible for inclusion (Figure 1).

**Figure 1:**
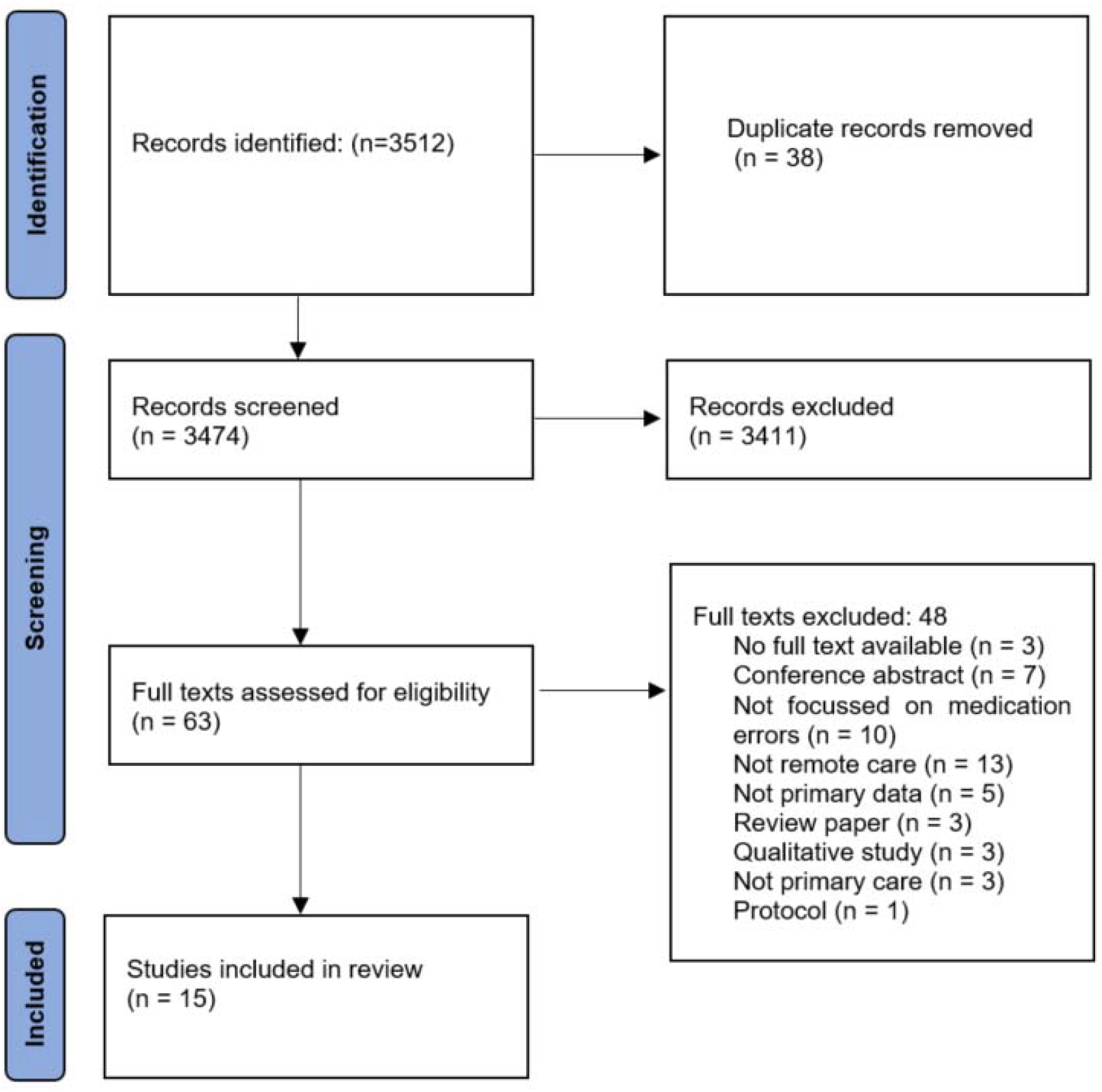
PRISMA Flow Diagram

### Study Characteristics

The characteristics of the included studies are detailed in Table 1. Of the 15 studies included in the review, 11 were conducted in the United States (US),(23–33) two in England,(34,35) one in Sweden,(36) and one in Finland.(37) All but two of the studies were carried out at multiple sites.(31,32) Eight cross-sectional studies with data collected prospectively were included,(24,26,29,31–34,36) along with two retrospective, cross-sectional studies,(23,25) one naturalistic stepped-wedge study,(35) one case series,(27) one controlled, before-and-after study,(28) one uncontrolled, before-and-after study,(30) and one cross-sectional survey of pharmacists’ opinions regarding the impact of e-prescriptions on medication safety.(37)

**Table 1:** Study Characteristics

| Study Author (Year) | Study Type | Setting & Country | Study Sample/Unit of Analysis | Outcome of Interest (n) | Data Collected By | Near Misses Reported | Incidents per 100 e-prescriptions Analysed |
| --- | --- | --- | --- | --- | --- | --- | --- |
| Åstrand et al (2009) (36) | Cross-sectional study (prospective data collection) | Three mail-order pharmacies in Sweden | 7532 e-prescriptions | Reasons for pharmacist intervention (n=150) | Independent researchers | No | 1.99 |
| Cochran et al (2013) (23) | Cross-sectional study (retrospective data collection) | Three pairs of ambulatory care clinics and community pharmacies in the mid-western United States | 403 e-prescriptions | Discrepancies between e-clinic instructions and pharmacy label (n=14) | Independent researchers | No | 3.47 |
| Franklin et al (2013) (34) | Cross-sectional study (prospective data collection) | Eight community pharmacies in England | n/a | Prescribing and dispensing errors (n=10) | Pharmacists/Tech nicians | No | n/a |
| Franklin et al (2014) (35) | Naturalistic stepped wedge study | Fifteen community pharmacies in England | 3733 items dispensed on e-prescriptions* | Dispensing errors (n=348) | Independent researchers | No | n/a |
| Gilligan et al (2012) (24) | Cross-sectional study (prospective data collection) | Two chain grocery store pharmacies located within the Phoenix metropolitan area (US) | 180 new e-prescriptions | Pharmacist interventions (n=21) | Independent researchers | No | 11.67 |
| Hincapie et al (2014) (26) | Cross-sectional study (prospective data collection) | Community pharmacies in the United States | 484 PEER Reports, 773 e-prescribing problems | Prescribing errors (n=400) | Pharmacists/Tech nicians | Yes | n/a |
| Hincapie et al (2019) (25) | Cross-sectional study (retrospective data collection) | Community pharmacies in the United States | 515 Pharmacy Provider e-prescribing Experience Reporting Portal (PEER) Reports, 550 Pharmacy Quality Commitment | Prescribing errors (n=1065) | Pharmacists/Tech nicians | Yes (PEER Reports only) | n/a |
| Kauppinen et al (2017) (37) | Cross-sectional survey | Community pharmacies in Finland | 143 pharmacists and 635 dispensers | Prescribing errors | Pharmacists/Tech nicians | No | n/a |
| Lourenco et al (2016) (27) | Case series | Community pharmacies in the United States | 5 medication incidents | Prescribing and dispensing errors (n=5) | Pharmacists/Tech nicians | No | n/a |
| Moniz et al (2011) (28) | Controlled, before-and-after study | Three clinics in the United States | 613 prescriptions | Prescribing and dispensing errors (n=16) | Independent researchers | No | 2.61 |
| Odukoya et al (2014) (29) | Cross-sectional study (prospective data collection) | Five community retail pharmacies in Southwest Wisconsin, US. | 45 observation hours | Prescribing and dispensing errors (n=75) | Independent researchers | No | n/a |
| Panich et al (2020) (30) | Uncontrolled, before-and-after study | Two outpatient pharmacies in the United States | 2017 e-prescriptions | Prescribing and dispensing errors (n=18) | Pharmacists/Tech nicians | No | 0.89 |
| Reed-Kane et al (2014) (31) | Cross-sectional study (prospective data collection) | One compounding pharmacy* in Arizona, US | 111 new e-prescriptions | Prescribing errors (n=91) | Pharmacists/Tech nicians | No | 81.98 |
| Reed-Kane et al (2014) (32) | Cross-sectional study (prospective data collection) | One compounding pharmacy* in Arizona, US | 138 e-prescriptions | Prescribing errors (n=32) | Pharmacists/Tech nicians | No | 23.19 |
| Warholak and Rupp (2008) (33) | Cross-sectional study (prospective data collection) | 68 chain community pharmacies in the United States | 2,690 e-prescription orders (new, 83.0%; refill, 17.0%) | Reasons for pharmacist intervention (n=113) | Pharmacists/Tech nicians | Yes | 4.20 |

US=United States, ICPS=International Classification for Patient Safety, QRE=Quality Related Event, Rx=Prescription
\*Compounding pharmacies specialise in preparing custom formulations of medications to meet unique patient needs.

Eleven studies investigated medication incidents identified in electronic prescriptions (e-prescriptions) in community pharmacies,(25–32,34,35,37) three investigated reasons for pharmacist interventions in e-prescriptions,(24,33,36) and one study investigated discrepancies between e-clinic instructions and the pharmacy label.(23) Although the focus of this review was medication safety incidents associated with both e-prescriptions and telemedicine, only studies investigating incidents associated with e-prescriptions were identified.

### Types and Frequency of Medication Safety Incidents

**Table 2** shows the incident types reported in each study, expressed as a percentage of all incidents. Although some studies focussed only limited types of errors (hence the predominance of particular errors in such cases), this table illustrates the relative weighting of different error types across studies. The most commonly identified incident types according to the ICPS were ‘wrong label/instruction’, reported in 11 studies,(23,25,26,28,29,31–36) ‘wrong dose/strength/frequency’, reported in 9 studies,(24–31,33) ‘wrong drug’, reported in 8 studies,(25–31,33) and ‘wrong quantity’, also reported in 8 studies.(36,24–31,33) ‘Wrong formulation’ incidents were reported in 5 studies, ‘wrong patient’ incidents in 4 studies, ‘contraindication’ incidents in 3 studies, and ‘wrong route’ incidents in one study. In three studies, more than half of the incidents reported were categorised as ‘Other’; examples of these incidents include incomplete prescriptions, missing prescriber address, or missing essential information.(30,34,36) The survey study by Kauppinen *et al*. reported that incorrect total amount of medication, missing notification of exceptional dosage instructions or exceptional purpose of use, and unclear or incorrect dosage instructions were perceived by survey respondents to be the most commonly occurring incident types.(37) No study reported medication safety incidents that fell into the following ICPS categories: ‘wrong storage’, ‘omitted medicine/dose’, ‘expired dose’, and ‘adverse drug reaction’.(22)

**Table 2:** Percentage of Errors with each WHO Taxonomy Error Type Reported in Each Study, with colouring indicating percentage within each study ranging from 0% (white) to 100% (red)*

| WHO Taxonomy | Åstrand et al (36) | Cochran et al (23) | Franklin et al (2013) (34) | Franklin et al (2014) (35) | Gilligan et al (24) | Hincapie et al (2014) (26) | Hincapie et al (2019) (25) | Lourenco et al (27) | Moniz et al (28) | Odukoya et al (29) | Panich et al (30) | Reed-Kane et al (2014a) (31) | Reed-Kane et al (2014b) (32) | Warholak and Rupp (33) |
| --- | --- | --- | --- | --- | --- | --- | --- | --- | --- | --- | --- | --- | --- | --- |
| Wrong label, instruction | 32 | 100 | 10 | 100 |  | 25.8 | 23.1 |  | 18.75 | 18.7 |  | 8.8 | 6.25 | 0.88 |
| Wrong dose, strength, frequency |  |  |  |  | 18.2 | 13.3 | 12.2 | 20 | 37.5 | 10.7 | 5.56 | 8.8 | 12.5 | 17.69 |
| Wrong drug |  |  |  |  |  | 13.3 | 13.6 | 40 |  | 4 | 5.56 | 5.5 | 12.5 | 2.65 |
| Wrong quantity | 5.3 |  |  |  | 27.3 | 15.8 | 11.7 | 20 |  | 61.3 | 5.56 |  |  | 10.61 |
| Wrong patient |  |  |  |  |  | 3.3 | 1.9 | 20 |  | 5.3 |  |  |  | 2.65 |
| Wrong formulation |  |  | 10 |  |  | 6.3 | 3.2 |  |  |  |  | 3.3 |  | 4.42 |
| Contraindication | 1.3 |  |  |  | 9.1 |  |  |  |  |  |  |  |  | 7.07 |
| Wrong route |  |  |  |  |  | 2.0 | 1.0 |  |  |  |  |  |  |  |
| QRE |  |  | 20 |  | 40.9 | 7.8 | 5.1 |  |  |  |  | 70.3 | 31.25 | 7.07 |
| Other | 61.3 |  | 60 |  | 4.5 | 12.8 | 28.2 |  | 43.75 |  | 83.33 | 3 | 37.5 | 46.9 |

QRE=Quality Related Event
\*WHO Taxonomy could not be applied to the results of Kauppinen et al.
\*\* No study reported events in the following categories: Wrong Storage, Omitted Medicine/Dose, Expired Medicine, Adverse Drug Reaction

There was significant heterogeneity across the identified studies regarding what was classed as a medication safety incident, and how these incidents were measured. For example, Åstrand *et al*. included ‘Incorrect quantity/duration of treatment’ as a single incident category, while Gilligan *et al*. reported ‘excessive quantity/duration’ and ‘insufficient quantity/duration’ as separate categories.(24,36) Furthermore, in six studies, the majority of incidents reported were categorised as either QREs or ‘other’. Over 70% of the incidents reported by Reed-Kane *et al*. were ‘wrong entry field errors’, when the drug name was written in the incorrect field in the e-prescribing software, which did not fall under the definition of medication safety incident used in this review.(31)

In six studies, medication safety incidents were documented by an independent researcher,(23,24,28,29,35,36) while in the remaining nine studies a pharmacist or member of the pharmacy team recorded incidents as they occurred.(25–27,30–34,37) Six studies reported prescribing errors only,(25,26,31–33,37) five studies reported both prescribing and dispensing errors,(27–30,34) two studies reported dispensing errors only,(23,35) and two reported reasons for pharmacist interventions in prescriptions.(24,36) The studies that reported both prescribing and dispensing errors did not specify at which stage in the medication process each error type occurred. Finally, just 3 of the 15 studies differentiated between medication errors and near misses.(25,26,33)

The frequency of medication safety incidents was reported or could be calculated for 8 of the 15 studies. As shown in **Table 1**, the frequency of medication safety incidents ranged from 0.89 to 81.98 incidents per 100 e-prescriptions analysed.

### Causes of Medication Safety Incidents

Three studies reported the causes of medication safety incidents associated with e-prescribing.(25,29,34) Franklin *et al*. reported the ‘origin’ of e-prescription problems; of the 10 problems reported in the study, 8 were clinical in origin (e.g. incomplete prescription) and 2 were due to organisational issues.(34) Hincapie *et al*. conducted a qualitative analysis of a sample of incident reports and concluded that human factors such as mistakes or miscalculations contributed to the majority of incidents, followed by communication and language barriers.(25) Through direct observations and interviews, Odukoya *et al*. identified three contributory factors: 1) incorrect calculation or entry of information, 2) auto-population of e-prescription information and 3) mismatch of e-prescription information between prescriber and pharmacy systems.(29)

### Outcomes of Medication Safety Incidents

Three studies reported the outcomes of medication safety incidents associated with e-prescribing.(25,27,29) The most commonly reported outcomes, each reported in two studies, were 1) incorrect medicine dispensed, 2) patient and pharmacist frustration, 3) slower or interrupted workflow, and 4) increased costs for patient and pharmacy.(25,29) Other reported outcomes were hospitalisation, adverse events and delayed therapy.(25,27,29)

### Relative Risk of Medication Safety Incidents

Five studies compared the rate of e-prescription incidents against the rate of incidents in traditional, paper-based prescriptions,(24,28,34–36) the results of which are displayed in **Table 3**. Four studies reported a measure of effect, (24,28,35,36) three of which found e-prescribing to be associated with higher levels of medication safety incidents than standard prescriptions.(24,35,36) Åstrand *et al*. found that e-prescriptions were significantly more likely to require a clarification contact than non-electronic prescriptions (RR 1.7 (95% CI 1.3-2.2)). Franklin *et al*. reported a statistically significant increase in labelling errors in e-prescriptions compared to standard prescriptions (OR 1.46 (95% CI 1.21 to 1.76)).(35) Gilligan *et al*. found that labelling errors were more likely to occur in e-prescriptions compared to standard prescriptions, however this finding was not statistically significant (RR 1.32 (95% CI 0.9-2.0), p<0.81).(24) Conversely, Moniz *et al*. reported a significant reduction in dispensing errors after the introduction of e-prescriptions (RR 0.5 (95% CI 0.3-0.8), p=0.034).(28)

**Table 3:** Studies with Comparison Groups

| Study Author | Year | Study Sample/Unit of Analysis | Comparator | Results | Measure of Effect |
| --- | --- | --- | --- | --- | --- |
| Åstrand *et al.* (2009) | 2009 | 7532 New e-prescriptions | 6833 New non-electronic prescriptions | Clarification contacts were made for 2.0% of new ePrescriptions and 1.2% of new non-electronic prescriptions. | RR 1.7 (95% CI 1.3-2.2) |
| Franklin *et al.* (2013) | 2013 | 69 'problems' identified in 68 e-prescriptions but total number of prescriptions analysed not reported | 59 'problems' associated with non-electronically transferred prescriptions | 0.51% of e-prescriptions contained an error compared with 0.40% of standard prescriptions. | n/a |
| Franklin *et al.* (2014) | 2014 | 3733 items dispensed on e-prescriptions (total number of e-prescriptions not reported) | 12,624 items dispensed on non-e-prescriptions | Labelling errors occurred in 7.4% of e-prescriptions compared with 4.8% of standard prescriptions. | OR 1.46 (95% CI 1.21 to 1.76) |
| Gilligan *et al.* (2012) | 2012 | 180 new e-prescriptions | 255 verbal, 561 fax and 682 handwritten Rx | Rates of intervention significantly higher in e-prescribing (11.7%) vs both faxed (3.9%) and verbal (5.1%) orders (P <.0001 and P=.008, respectively). E-prescribing (11.7%) vs handwritten (15.4%) prescriptions not significant (P<.21). Rate of e-prescribing interventions not significant compared with overall intervention rate (11.7% vs 9.4%;P <.81) | RR 1.32 (95% CI 0.9-2.0) |
| Moniz *et al.* (2011) | 2011 | 613 prescriptions | Control clinics (pre/post intervention without ET), E-prescribing clinic (pre intervention without ET, post intervention with ET) | Reduction in dispensing errors from 3.1% of prescriptions in the baseline period in e-prescribing clinic to 1.8% after the introduction of ET, p=0.034. No change among control clinics between pre and post intervention. | RR 0.5 (95% CI 0.3-0.8) |

ET=Electronic
Transfer

### Quality of Included Studies

The results of the quality assessment of the included studies are presented in **Table 4**. All included studies were found to be of acceptable methodological quality, although Åstrand *et al*. and Gilligan *et al*., which analysed the difference in rate/risk of errors/incidents, did not consider confounders in their analysis.(24,36)

**Table 4:** Quality Assessments using the CASP Checklist for cohort studies

| Study Author | Year | Did the study address a clearly focussed issue? | Was the cohort recruited in an acceptable way? | Was the exposure accurately measured to minimise bias? | Was the outcome accurately measured to minimise bias? | Have the authors identified all confounding factors? | Have they taken account of confounding factors in the design/analysis? | Was the follow up of subjects complete enough? | Was the follow up of subjects long enough? | How precise are the results? | Do you believe the results? | Can the results be applied to the local population? | Do the results of the study fit with the other available evidence? |
| --- | --- | --- | --- | --- | --- | --- | --- | --- | --- | --- | --- | --- | --- |
| Åstrand *et al.* | 2009 | Yes | Yes | Yes | Yes | No | No | n/a | n/a | Precise | Yes | Can't tell | Yes |
| Cochran *et al.* | 2013 | Yes | Yes | Yes | Yes | n/a | n/a | n/a | n/a | n/a | Yes | Yes | Yes |
| Franklin *et al.* | 2013 | Yes | Yes | Yes | Yes | n/a | n/a | n/a | n/a | n/a | Yes | Yes | Yes |
| Franklin *et al.* | 2014 | Yes | Yes | Yes | Yes | Yes | Yes | n/a | n/a | Precise | Yes | Yes | Yes |
| Gilligan *et al.* | 2012 | Yes | Yes | Yes | Yes | Can't tell | No | Can't tell | n/a | n/a | Yes | Yes | Yes |
| Hincapie *et al.* | 2019 | Yes | Yes | Yes | n/a | n/a | n/a | n/a | n/a | n/a | Yes | Yes | Yes |
| Hincapie *et al.* | 2014 | Yes | Yes | Yes | n/a | n/a | n/a | n/a | n/a | n/a | Yes | Yes | Yes |
| Kauppinen *et al.* | 2017 | Yes | Yes | Yes | n/a | n/a | n/a | n/a | n/a | n/a | Yes | Yes | Yes |
| Lourenco *et al.* | 2016 | Yes | Yes | Yes | n/a | n/a | n/a | n/a | n/a | n/a | Yes | Yes | Yes |
| Moniz *et al.* | 2011 | Yes | Yes | Yes | Yes | n/a | n/a | n/a | n/a | n/a | Yes | Yes | Yes |
| Odukoya *et al.* | 2014 | Yes | Yes | Yes | Yes | n/a | n/a | n/a | n/a | n/a | Yes | Yes | Yes |
| Panich *et al.* | 2020 | Yes | Yes | Yes | Yes | n/a | n/a | n/a | n/a | n/a | Yes | Yes | Yes |
| Reed-Kane *et al.* | 2014 | Yes | Yes | Yes | Can't tell | n/a | n/a | n/a | n/a | n/a | Yes | Yes | Yes |
| Reed-Kane *et al.* | 2014 | Yes | Yes | Yes | Can't tell | n/a | n/a | n/a | n/a | n/a | Yes | Yes | Yes |
| Warholak *et al.* | 2008 | Yes | Yes | Yes | Yes | n/a | n/a | n/a | n/a | n/a | Yes | Yes | Yes |

## Discussion

### Summary of Findings

The purpose of this review was to identify and examine the available evidence on medication safety incidents associated with the remote delivery of primary care, with a focus on electronic prescribing (e-prescribing) and telemedicine. Fifteen studies were identified that investigated medication safety incidents associated with e-prescribing, however no studies investigating medication safety incidents associated with telemedicine were identified. Due to the increased use of telemedicine since the beginning of the COVID-19 pandemic, this highlights an important gap in the literature.

Regarding the primary outcome of this review, the most common incident type reported in community pharmacies was ‘wrong label/instruction’, followed by ‘wrong dose/strength/frequency’, ‘wrong drug’ and ‘wrong quantity’. There is limited published literature on the types, causes and outcomes of medication safety incidents associated with the standard delivery of primary care.(38) Nonetheless, the results of this review could be compared to those of a 2009 systematic review which found that the most common dispensing errors in community pharmacies were supply of the wrong drug, strength, quantity or form and printing the wrong directions on the label.(39)

The secondary outcomes of this review were the frequency, causes, and outcomes of medication safety incidents associated with remote care. Variation in the identified studies and the lack of published data on rates of medication safety incidents in primary care make it difficult to determine whether e-prescriptions are associated with a higher frequency of medication safety incidents than standard prescriptions. According to the studies included in this review, between 0.89 and 81.98 incidents occurred for every 100 e-prescriptions analysed. This wide range of frequency estimates can be compared to a systematic review by Assiri *et al*., which found that the reported prevalence of prescribing errors in primary care ranged from 2% to 94%.(15) In that review, a broad definition of prescribing error was used, including potentially inappropriate prescribing and the prescribing of medications without a recorded indication. Similarly, due to the small number of studies that reported them, and the variation across these studies in terms of the e-prescribing systems used and how they were implemented, it is difficult to draw any conclusions regarding the causes and outcomes, including patient harm, of medication safety incidents associated with e-prescribing.

A significant finding of this review was the heterogeneity of the available evidence on medication safety incidents associated with e-prescribing. The studies included in this review varied greatly in terms of the definition and categories of medication safety incidents used, how incidents were reported and measured, who reported medication safety incidents, and even the definitions of e-prescribing used. In order to fully understand whether there is a medication safety risk associated with e-prescribing, and the extent of that risk, better reporting standards are required. However, the issue of poor reporting standards is not restricted to remote care or even primary care. Across health services research there is a lack of consensus regarding how medication safety incidents and near misses should be defined and categorised.(40) A key principle of patient safety research is that medication safety incidents must be reported in order to learn from errors and prevent them from happening again.(41) The ICPS used to categorise medication safety incidents in this study is not widely applied in the literature, despite being made available by the WHO over a decade ago.(42) Furthermore, with the increasing use of e-prescribing and telemedicine worldwide, a revised version of the ICPS is potentially required to reflect the emergence of new incident types related to remote care, as evidenced by the high percentage of incidents categorised as ‘Other’ in this review. Until an updated, standardised system for reporting medication safety incidents is applied by healthcare professionals and researchers worldwide, the goal of ‘Medication Without Harm’ will likely remain out of reach.(9)

### Implications for Practice

To the best of our knowledge, this is the first review to examine the evidence on the types and frequencies of medication safety incidents associated with the remote delivery of primary care. The findings of this review are relevant for countries where e-prescribing has been in place for many years, however, the impact of this review will likely be most important in countries such as Ireland, Austria and Italy in Europe, where the electronic transfer of prescriptions was introduced in response to the COVID-19 pandemic, or countries where the introduction of e-prescribing or electronic prescription transfer is under consideration.(3,6)

Electronic transfer of prescriptions has been widely used in countries such as Sweden, England, Scotland, the US, Denmark and the Netherlands for several years, and countries that have introduced integrated e-prescribing systems have benefitted from safer and more streamlined medicines management processes.(43–45) In Ireland, legislation for electronic prescription transfer was only introduced in April 2020, as part of a series of countermeasures to prevent the spread of COVID-19 in healthcare facilities.(6) In contrast with countries such as England and the Netherlands, that spent years developing an integrated e-prescribing system, the switch to electronic prescription transfer occurred very rapidly in Ireland, with little to no training or integration into existing systems.(6,20)

In their review, Odukoya *et al*. identified a mismatch of e-prescription information between prescriber and pharmacy systems as a potential cause of medication safety incidents.(29) The current system in Ireland is not an integrated e-prescribing system but a secure email service to support the electronic transfer of prescriptions; the lack of an integrated system could potentially increase the risk of medication safety incidents. Also, because the current interim system was implemented more rapidly in Ireland than other countries, it is possible that the risk and types of incidents may differ in this context, compared to other settings where e-prescribing systems were thoroughly integrated. It would therefore be pertinent for Irish healthcare policymakers to collaborate with academics, GPs and community pharmacists to evaluate the current interim electronic prescription transfer system and investigate how it could be improved.

While it is too early yet to determine the effects of COVID-19 prevention measures on medication safety in primary care, it is clear that the pandemic has had a profound impact on the way healthcare professionals and patients engage with and experience primary care services.(3,4) As the COVID-19 pandemic evolves, changes such as the use of telemedicine and e-prescribing could play a role in the provision of primary care for the foreseeable future. It is imperative that any strategies put in place to protect patients during the pandemic, from e-prescribing to changes in prescribing guidelines, are thoroughly evaluated and implemented with medication safety best practice in mind.

### Strengths and Limitations

Due to the emerging nature of the review topic, a rapid review was carried out which, according to the Cochrane Rapid Reviews Methods Group, *‘accelerates the process of conducting a traditional systematic review through streamlining or omitting specific methods to produce evidence for stakeholders in a resource-efficient manner’*.(17) A strength of this review was the timely provision of high-quality evidence. This rapid review followed guidance from the Cochrane Rapid Reviews Methods Group and included a search strategy developed with input from a wide group of experts, including a librarian and a systematic reviews expert. Although study selection, data abstraction and risk of bias assessment performed were not performed in duplicate, each step was verified by a second reviewer. However, as with any review, there were also some limitations. Results were limited to English language studies published since the year 2000, and the search was limited to two online databases. The majority of the included studies were conducted in the US, which could limit the generalisability of the review findings. Finally, Ireland has implemented a system for the electronic transfer of prescriptions, as opposed to a comprehensive e-prescribing system, therefore the findings of this study may not accurately reflect the issues currently encountered in the Irish healthcare system.

### Further Research

This review has highlighted a number of important areas for further research. A gap in the literature was identified regarded the impact of telemedicine on medication safety. As telemedicine becomes more widely used, research should be conducted on its safety and efficacy, and on the experiences of healthcare providers and patients delivering and receiving care remotely. The identified studies on medication safety incidents associated with e-prescribing varied widely in terms of what was reported as a medication safety incident and how such incidents were recorded. Future research in this field should use standardised methods and definitions to allow for comparisons between studies. As the COVID-19 pandemic continues to evolve, it is imperative that changes to primary care are evaluated and that research is conducted on the impact of the pandemic on medication safety and patient outcomes. Finally, as mentioned previously, the findings of this review will be used to inform the design of a toolkit to support safe medication use during COVID-19.

## Conclusions

To the best of our knowledge, this was the first review to examine the literature on medication safety incidents associated with the remote delivery of primary care. A lack of published evidence on the safety of telemedicine was highlighted. The included studies that investigated medication safety incidents associated with e-prescribing varied in terms of how incidents were recorded and categorised. The most commonly reported medication safety incident types associated with e-prescribing were ‘wrong label/instruction’ and ‘wrong dose/strength/frequency’. The frequency of medication safety incidents ranged from 0.89 to 81.98 incidents per 100 e-prescriptions analysed. Further research is needed to evaluate the safety and efficacy of e-prescribing and telemedicine, and the impact of the COVID-19 pandemic on medication safety.

## Data Availability

All data extracted from papers as part of this rapid review are contained within the manuscript.

## Appendix 1

### PRISMA Checklist

| Section and Topic | Item # | Checklist item | Location where item is reported |
| --- | --- | --- | --- |
| **TITLE** |  |  |  |
| Title | 1 | Identify the report as a systematic review. | 1 |
| **ABSTRACT** |  |  |  |
| Abstract | 2 | See the PRISMA 2020 for Abstracts checklist. | 2 |
| **INTRODUCTION** |  |  |  |
| Rationale | 3 | Describe the rationale for the review in the context of existing knowledge. | 4-5 |
| Objectives | 4 | Provide an explicit statement of the objective(s) or question(s) the review addresses. | 5 |
| **METHODS** |  |  |  |
| Eligibility criteria | 5 | Specify the inclusion and exclusion criteria for the review and how studies were grouped for the syntheses. | 6-7 |
| Information sources | 6 | Specify all databases, registers, websites, organisations, reference lists and other sources searched or consulted to identify studies. Specify the date when each source was last searched or consulted. | 5-6 |
| Search strategy | 7 | Present the full search strategies for all databases, registers and websites, including any filters and limits used. | 33-34 |
| Selection process | 8 | Specify the methods used to decide whether a study met the inclusion criteria of the review, including how many reviewers screened each record and each report retrieved, whether they worked independently, and if applicable, details of automation tools used in the process. | 5-7 |
| Data collection process | 9 | Specify the methods used to collect data from reports, including how many reviewers collected data from each report, whether they worked independently, any processes for obtaining or confirming data from study investigators, and if applicable, details of automation tools used in the process. | 7-8 |
| Data items | 10a | List and define all outcomes for which data were sought. Specify whether all results that were compatible with each outcome domain in each study were sought (e.g. for all measures, time points, analyses), and if not, the methods used to decide which results to collect. | 7-8 |
|  | 10b | List and define all other variables for which data were sought (e.g. participant and intervention characteristics, funding sources). Describe any assumptions made about any missing or unclear information. | 7-8 |
| Study risk of bias assessment | 11 | Specify the methods used to assess risk of bias in the included studies, including details of the tool(s) used, how many reviewers assessed each study and whether they worked independently, and if applicable, details of automation tools used in the process. | 8 |
| Effect measures | 12 | Specify for each outcome the effect measure(s) (e.g. risk ratio, mean difference) used in the synthesis or presentation of results. | 8 |
| Synthesis | 13a | Describe the processes used to decide which studies were eligible for each synthesis (e.g. tabulating the study intervention characteristics and | 8 |
| methods |  | comparing against the planned groups for each synthesis (item #5)). |  |
|  | 13b | Describe any methods required to prepare the data for presentation or synthesis, such as handling of missing summary statistics, or data conversions. | 8 |
|  | 13c | Describe any methods used to tabulate or visually display results of individual studies and syntheses. | 8 |
|  | 13d | Describe any methods used to synthesize results and provide a rationale for the choice(s). If meta-analysis was performed, describe the model(s), method(s) to identify the presence and extent of statistical heterogeneity, and software package(s) used. | n/a |
|  | 13e | Describe any methods used to explore possible causes of heterogeneity among study results (e.g. subgroup analysis, meta-regression). | n/a |
|  | 13f | Describe any sensitivity analyses conducted to assess robustness of the synthesized results. | n/a |
| Reporting bias assessment | 14 | Describe any methods used to assess risk of bias due to missing results in a synthesis (arising from reporting biases). | n/a |
| Certainty assessment | 15 | Describe any methods used to assess certainty (or confidence) in the body of evidence for an outcome. | n/a |
| **RESULTS** |  |  |  |
| Study selection | 16a | Describe the results of the search and selection process, from the number of records identified in the search to the number of studies included in the review, ideally using a flow diagram. | 8-9 |
|  | 16b | Cite studies that might appear to meet the inclusion criteria, but which were excluded, and explain why they were excluded. | n/a |
| Study characteristics | 17 | Cite each included study and present its characteristics. | 8-12 |
| Risk of bias in studies | 18 | Present assessments of risk of bias for each included study. | 18-19 |
| Results of individual studies | 19 | For all outcomes, present, for each study: (a) summary statistics for each group (where appropriate) and (b) an effect estimate and its precision (e.g. confidence/credible interval), ideally using structured tables or plots. | 12-17 |
| Results of syntheses | 20a | For each synthesis, briefly summarise the characteristics and risk of bias among contributing studies. | 18-19 |
|  | 20b | Present results of all statistical syntheses conducted. If meta-analysis was done, present for each the summary estimate and its precision (e.g. confidence/credible interval) and measures of statistical heterogeneity. If comparing groups, describe the direction of the effect. | 12-17 |
|  | 20c | Present results of all investigations of possible causes of heterogeneity among study results. | n/a |
|  | 20d | Present results of all sensitivity analyses conducted to assess the robustness of the synthesized results. | n/a |
| Reporting biases | 21 | Present assessments of risk of bias due to missing results (arising from reporting biases) for each synthesis assessed. | n/a |
| Certainty of | 22 | Present assessments of certainty (or confidence) in the body of evidence for each outcome assessed. | n/a |
| evidence |  |  |  |
| **DISCUSSION** |  |  |  |
| Discussion | 23a | Provide a general interpretation of the results in the context of other evidence. | 21-25 |
|  | 23b | Discuss any limitations of the evidence included in the review. | 21-25 |
|  | 23c | Discuss any limitations of the review processes used. | 21-25 |
|  | 23d | Discuss implications of the results for practice, policy, and future research. | 21-25 |
| **OTHER INFORMATION** |  |  |  |
| Registration and protocol | 24a | Provide registration information for the review, including register name and registration number, or state that the review was not registered. | 5 |
|  | 24b | Indicate where the review protocol can be accessed, or state that a protocol was not prepared. | 5 |
|  | 24c | Describe and explain any amendments to information provided at registration or in the protocol. | n/a |
| Support | 25 | Describe sources of financial or non-financial support for the review, and the role of the funders or sponsors in the review. | n/a |
| Competing interests | 26 | Declare any competing interests of review authors. | n/a |
| Availability of data, code and other materials | 27 | Report which of the following are publicly available and where they can be found: template data collection forms; data extracted from included studies; data used for all analyses; analytic code; any other materials used in the review. | n/a |

## Appendix 2

### Search Strategy – 02/06/21

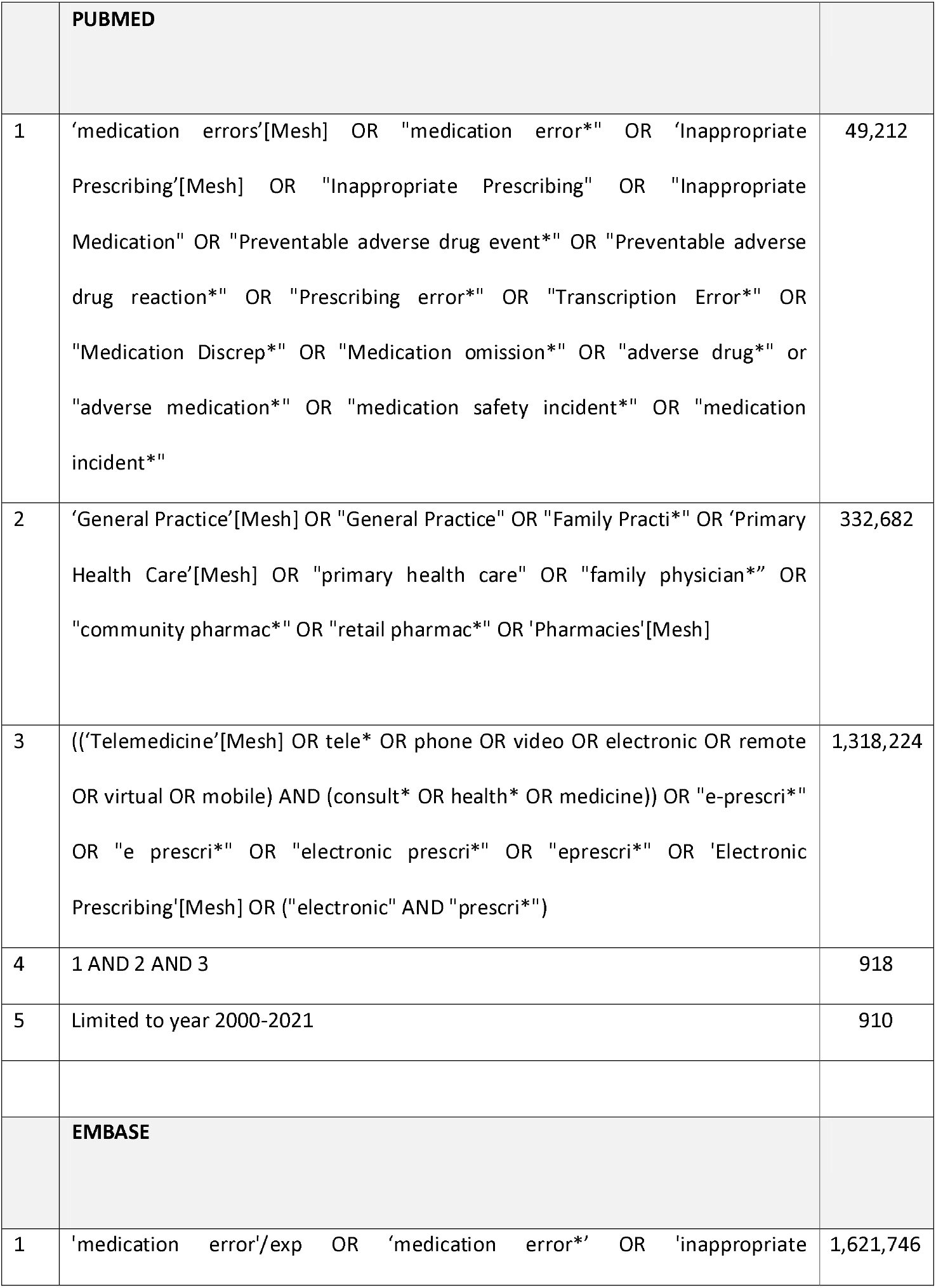
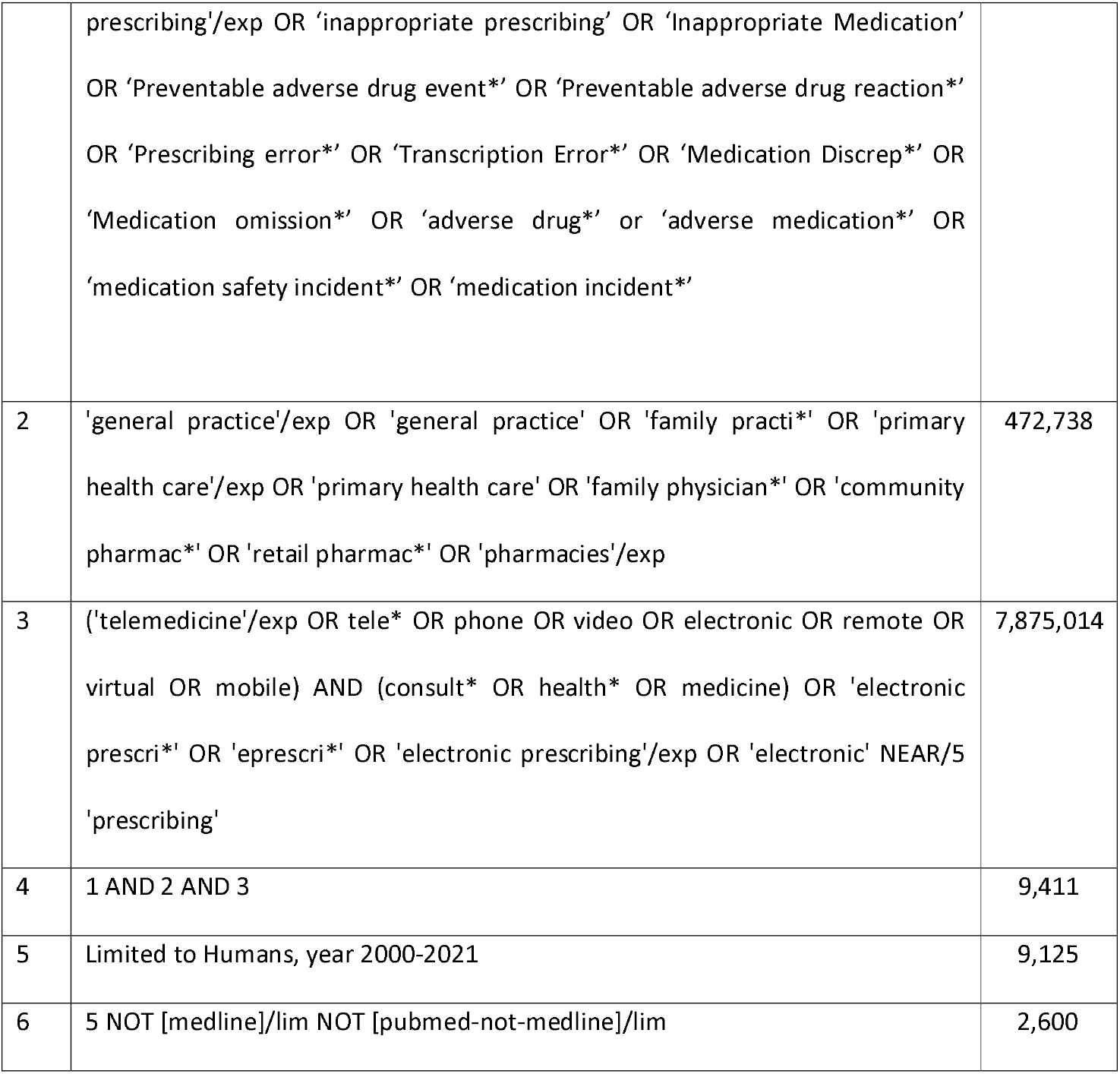

|  |  |  |
| --- | --- | --- |
|  | **PUBMED** |  |
| 1 | 'medication errors'[Mesh] OR "medication error*" OR 'Inappropriate Prescribing'[Mesh] OR "Inappropriate Prescribing" OR "Inappropriate Medication" OR "Preventable adverse drug event*" OR "Preventable adverse drug reaction*" OR "Prescribing error*" OR "Transcription Error*" OR "Medication Discrep*" OR "Medication omission*" OR "adverse drug*" or "adverse medication*" OR "medication safety incident*" OR "medication incident*" | 49,212 |
| 2 | 'General Practice'[Mesh] OR "General Practice" OR "Family Practi*" OR 'Primary Health Care'[Mesh] OR "primary health care" OR "family physician*" OR "community pharmac*" OR "retail pharmac*" OR 'Pharmacies'[Mesh] | 332,682 |
| 3 | ((('Telemedicine'[Mesh] OR tele* OR phone OR video OR electronic OR remote OR virtual OR mobile) AND (consult* OR health* OR medicine)) OR "e-prescri*" OR "e prescri*" OR "electronic prescri*" OR "eprescri*" OR 'Electronic Prescribing'[Mesh] OR ("electronic" AND "prescri*")) | 1,318,224 |
| 4 | 1 AND 2 AND 3 | 918 |
| 5 | Limited to year 2000-2021 | 910 |
|  | **EMBASE** |  |
| 1 | 'medication error'/exp OR 'medication error*' OR 'inappropriate | 1,621,746 |
|  | prescribing'/exp OR 'inappropriate prescribing' OR 'Inappropriate Medication'<br>OR 'Preventable adverse drug event*' OR 'Preventable adverse drug reaction*'<br>OR 'Prescribing error*' OR 'Transcription Error*' OR 'Medication Discrep*' OR<br>'Medication omission*' OR 'adverse drug*' or 'adverse medication*' OR<br>'medication safety incident*' OR 'medication incident*' |  |
| 2 | 'general practice'/exp OR 'general practice' OR 'family practi*' OR 'primary<br>health care'/exp OR 'primary health care' OR 'family physician*' OR 'community<br>pharmac*' OR 'retail pharmac*' OR 'pharmacies'/exp | 472,738 |
| 3 | ('telemedicine'/exp OR tele* OR phone OR video OR electronic OR remote OR<br>virtual OR mobile) AND (consult* OR health* OR medicine) OR 'electronic<br>prescri*' OR 'eprescri*' OR 'electronic prescribing'/exp OR 'electronic' NEAR/5<br>'prescribing' | 7,875,014 |
| 4 | 1 AND 2 AND 3 | 9,411 |
| 5 | Limited to Humans, year 2000-2021 | 9,125 |
| 6 | 5 NOT [medline]/lim NOT [pubmed-not-medline]/lim | 2,600 |

